# Automating clinical trial outcome identification and misreporting detection using RegCheck

**DOI:** 10.64898/2026.07.30.26358885

**Authors:** Jamie Cummins, Henry Drysdale, Malte Elson, Ian Hussey, Ben Goldacre, Nicholas J. DeVito

## Abstract

**Objective:** To evaluate the accuracy and cost of RegCheck, an automated large language model (LLM)-based workflow, for identifying clinical trial outcomes and detecting outcome misreporting by comparing its outputs with manual assessments from the COMPare Trials project.

**Design:** Validation study.

**Setting:** Sixty-two clinical trials originally assessed in the COMPare Trials project, sampled from five high impact general medical journals.

**Participants:** Published clinical trial reports and their corresponding prespecified registrations and/or protocols.

**Main outcome measures:** Four prespecified research questions were examined. RQ1 assessed outcome extraction recall relative to COMPare. RQ2 assessed accuracy of outcome classification as primary, secondary, or non-prespecified. RQ3 assessed accuracy of misreporting detection relative to COMPare, with additional manual adjudication of discrepancies between RegCheck and COMPare. RQ4 assessed the average per-paper cost of running the automated workflow.

**Results:** Across the validation papers, RegCheck achieved 91.2% outcome extraction recall relative to COMPare, and 83.6% outcome classification accuracy. For detection of outcome misreporting, RegCheck’s overall accuracy was 85.6%. However, after resolving discrepancies with the original human judgements (which frequently favoured RegCheck’s judgement), revised accuracy for outcome misreporting detection was 94.8%. The mean cost of running the workflow was 5.94 USD per paper.

**Conclusions:** RegCheck achieved high overall performance with a rigorous manual benchmark for identifying prespecified and reported outcomes in clinical trials, and detecting outcome misreporting, while operating at very low marginal cost. Adjudication of discrepant judgements suggested that RegCheck frequently identified valid issues not captured in the reference standard. Automated outcome checking may offer a scalable way to support editors, peer reviewers, and authors in detecting outcome switching and improving trial reporting.

**What is already known on this topic:** Undeclared discrepancies between prespecified and reported trial outcomes remain common and can distort the medical evidence base.

Manual auditing projects such as COMPare Trials have shown that outcome switching is prevalent, but this work is labour intensive and difficult to sustain at scale.

Large language models may help automate registration-to-publication comparisons, but their performance against established manual benchmarks has not been well characterised.

**What this study adds:** RegCheck, an LLM-based workflow, achieved high performance for outcome extraction, generally high performance for outcome classification (with three protocols exhibiting exceptionally poor performance), and high performance for misreporting detection when benchmarked against COMPare Trials.

Manual adjudication of disagreements in misreporting judgements suggested that RegCheck’s judgements of misreporting were (at times) more accurate than those of the human reports.

The workflow operated at a low mean cost per paper, suggesting that automated trial checking could be deployed at scale as a screening and decision-support tool.

## Introduction

Selective and inaccurate reporting of outcomes remains a major threat to the credibility of clinical trial evidence (1,2). Undeclared discrepancies between prespecified and reported outcomes can inflate apparent treatment benefits, obscure null or adverse findings, and compromise evidence synthesis and clinical decision-making (3). Best-practice guidance, including CONSORT and related methodological standards, requires transparent reporting of prespecified outcomes and clear disclosure of any changes made after trial commencement (4,5). Yet substantial evidence indicates that discrepant reporting remains common across the medical literature (6,7).

The COMPare Trials project provided one of the clearest demonstrations of the scale of this problem. In this project, Goldacre and colleagues manually compared study registrations/protocols to corresponding published papers. Specifically, they examined the accuracy of outcome-reporting in all trials published across a six week period in five leading medical journals which endorsed the CONSORT guidelines (8). They found that undeclared discrepancies were widespread, with 87% of trials exhibiting problems in this regard. The authors found that when they contacted those journals regarding these issues, their post-publication correction efforts were often met with limited engagement. This work was methodologically influential, but it also highlighted a core practical constraint: high quality discrepancy checking is slow, effortful, and difficult to sustain using human labour alone. Indeed, Goldacre and colleagues noted as much themselves, stating that “the workload associated with checking trials in real time, in detail, and maintaining subsequent interactive correspondence within the timeline for publication was extremely high, even for a large coordinated team” (8).

Trial registration was introduced partly to reduce publication bias and partly to deter undeclared outcome switching by creating a time-stamped public record of prespecified plans (9). But registration alone cannot prevent selective reporting – it merely creates the possibility of detection. For that possibility to become an effective safeguard, the registry/protocol and the published paper must actually be compared by someone. Without this, discrepancies will simply not be identified (2). In practice, such scrutiny is infrequently done, under-resourced, and rarely incentivized (10,11).

Recent advances in natural language processing and generative large language models (LLMs) create a new opportunity to address this scalability problem. LLMs can extract, compare, and summarise complex and unstructured textual information across multiple documents. Recently, some of the current authorship team have developed RegCheck, an open-source software tool designed to extract relevant text from study registrations/protocols and published papers, compare them, and evaluate the consistency of these materials (12). In principle, such an LLM-based workflow could provide rapid first-pass checks of outcome reporting accuracy, flagging likely discrepancies for further review by editors, reviewers, or authors. Of course, such a workflow would be useful only if it achieved sufficiently high accuracy relative to careful human assessment.

In this study, we developed and evaluated a variant of RegCheck which was aimed specifically at the extraction of outcomes from study registrations/protocols and papers, and the comparison of the consistency of reporting between these materials in accordance with CONSORT. To do this, we used the COMPare Trials dataset as an “alloyed gold standard” (i.e., a reference standard we expect has errors; see 13) and examined the extent to which RegCheck’s output concorded with the original COMPare reports. We focused on four research questions: how accurately RegCheck extracted outcomes; how accurately it classified outcomes as primary, secondary, or non-prespecified; how accurately it detected misreporting; and how much it cost to run per paper.

## Methods

### Study Design

We conducted a validation study comparing outputs from RegCheck with manual assessments from the COMPare Trials project. The study followed a registered analysis plan, with COMPare treated as an alloyed gold standard. Where discrepancies arose between the automated workflow and COMPare, these were manually reviewed and annotated by the authorship team.

### Sample

The original COMPare Trials project examined 67 clinical trials. As registered, we used five trials as material for the workflow development of the RegCheck pipeline (Reports 5, 6, 35, 44, and 45), leaving a validation sample of 62 trials for the main analysis. After exclusions due to a small number of issues (detailed below) this left us with a final sample of 59 clinical trials for analysis.

For each trial, we used the same materials for comparison as were used by the original COMPare report. In other words: if the COMPare report had used the study registration as the main source of prespecification, so did we; if the COMPare report had used the study protocol as the main source of prespecification, so did we. Where multiple registration or protocol versions existed, we used the version that had also been used in the production of the COMPare report.

### RegCheck Workflow

The RegCheck workflow consisted of seven stages. First, registration or protocol text was ingested. ClinicalTrials.gov records were obtained through the registry API and converted from JSON to a readable text representation. Other documents were parsed as text using the DPT-2 document parsing model from LandingAI. Second, the published trial report was ingested from the journal PDF using GROBID: a machine learning library specialized for academic text extraction (14). Third, both prespecification and publication documents were prepared for retrieval-augmented generation: documents were segmented into chunks, semantic embeddings were computed using the text-embedding-3-large model from OpenAI, and the most relevant excerpts were retrieved for downstream queries (15).

Fourth, all distinct outcomes from the registration or protocol were extracted using an LLM (gpt-5.1-2025-11-13 on high reasoning effort). Outcomes were defined in accordance with COMPare’s protocol, requiring the specification of the variable measured, the method of measurement, and the time frame (8). Fifth, each extracted prespecified outcome was classified as primary, secondary, or missing/unspecified based on the registration or protocol text, once again using distinct calls to the LLM. Sixth, each prespecified outcome was aligned to the paper, and the paper was assessed for whether the outcome was reported, how it was designated in the manuscript, and whether reporting was accurate under CONSORT items 6a and 6b. Finally, paper-only outcomes were identified and evaluated based on whether they were transparently reported as non-prespecified.

Full prompts are reported in the study registration and are available in the supplementary materials. The code for RegCheck is open source and the specific implementation for outcome misreporting detection as used in the current study is available at https://osf.io/mxacq.

### Development and Piloting

The workflow was developed and refined on the five randomly-selected COMPare trials mentioned above. These papers were used to adapt the architecture for the clinical trial use case, to build the analysis pipeline, and catch any oddities or idiosyncrasies that were noted for this specific type of pipeline design. This also enabled us to ensure that certain ingestion features of the pipeline (e.g., only ingesting ClinicalTrials.gov registry entries made before the date recruitment had started) were interfacing correctly with the LLM-based portions of the workflow. These five pilot trials were excluded from validation analyses.

### Outcome Harmonisation

Because the wording of outcomes extracted by RegCheck and by COMPare would likely differ while referring to the same underlying outcome (in the same manner that extractions from different humans would also differ from one another), we manually harmonised the two outputs into a combined dataset after the RegCheck reports were generated. Each RegCheck outcome was reviewed alongside the corresponding COMPare outcome so that they could be directly compared in our analyses. In some cases, extractions in the original COMPare reports pooled outcomes together in a single row and evaluated them in aggregate (e.g., “EuroQol overall score and each subscale score” may be extracted as one outcome) whereas RegCheck decomposed those into distinct sub-outcomes in line with CONSORT-style specificity and provided separate evaluations for each of these (e.g., outcomes would be extracted as: 1. EuroQol overall score, 2. EuroQol mobility subscale, …). For these cases, we prespecified the following amalgamation rule: these distinct RegCheck outcomes were treated as a single comparison unit against the single unit from COMPare, and the most binding classification or reporting judgment was used where RegCheck’s evaluations of sub-outcomes disagreed. In other words: if a single suboutcome were coded as primary while the remaining were coded as secondary, then the concatenated outcome would be coded as primary; if a single suboutcome were coded as incorrectly reported while the remaining were coded as correctly reported, then the concatenated outcome would be coded as incorrectly reported.

### Outcomes

Our four prespecified research questions were as follows:

**RQ1**: how accurately did RegCheck extract outcomes relative to COMPare?

**RQ2**: how accurately did RegCheck classify extracted outcomes as primary, secondary, or non-prespecified?

**RQ3**: how accurately did RegCheck identify outcome misreporting relative to COMPare?

**RQ4**: what was the average cost of running the workflow per paper?

### Statistical Analysis

Unless otherwise specified, all estimates are reported with 95% confidence intervals obtained from cluster bootstrapping at the paper-level. For RQ1, we estimated the proportion of COMPare outcomes that were also identified by RegCheck. In our registration this was framed as recall relative to COMPare; for readability we refer to it here as extraction accuracy relative to the reference standard. For RQ2, we analysed only outcomes present in both COMPare and RegCheck, because classification cannot be assessed for missed outcomes. We compared RegCheck classification accuracy relative to the COMPare classifications across three categories: primary, secondary, and non-prespecified.

For RQ3, we assessed binary judgments of accurate reporting versus misreporting. In keeping with our registration, this analysis was restricted to outcomes that had been correctly identified (i.e., RQ1) and appropriately classified (i.e., RQ2). We additionally manually reviewed discrepancies between RegCheck and COMPare to assess the number of cases which were the result of inaccuracies on the part of RegCheck, or on the part of the original human-based judgements^1^. For RQ4, we calculated the average cost per paper by dividing the total workflow cost by the number of analysed papers.

### Patient and Public Involvement

Patients and members of the public were not involved in the design, conduct, reporting, or dissemination plans of this methodological validation study.

## Results

### Sample Characteristics

The validation sample comprised the 62 trial reports drawn from COMPare Trials which were not used in our piloting and development. Issues with four trials in the validation sample arose while running the workflow. Specifically, Study 1 from Trial 12 was removed because the protocol document could not be ingested successfully. The protocol from Study 2 from Trial 12 was ingested successfully. Therefore, Trial 12 remained included but only with a focus on those outcomes in Study 2. Separately, the paper for Trial 32 was in a Letter format, which caused issues in ingesting it via GROBID (which expects papers to be formatted in a traditional academic paper format). Trial 32 was thus excluded from when only disagreeing cases are inspected. We are currently in the process of a manual audit of a sample of agreeing cases, which we will also factor into this revised estimate. This analysis will be completed, and this working paper’s figures updated, prior to submission of this work to a peer-reviewed journal. Readers should note in any case that our registered primary outcome for RQ3 is the pre-adjudicated accuracy, and this post-adjudicated accuracy should thus be interpreted cautiously as an additional unregistered outcome in any case. analyses. Additionally, two trials in the original COMPare study (Trial 34 and Trial 43) were not compared against a registration or protocol, as all versions of the registration/protocol were registered after the start of participant recruitment. We therefore also excluded these two trials from our analyses. This meant that our final, analysed sample consisted of a total of 59 clinical trials. Across these trials, prespecification sources varied as either being based on the protocol or the study registration, in line with the source used in the original COMPare study. Specifically, 40 reports were based on the study registrations, and 19 reports were based on the study protocol. A summary of the main results of this work is reported in Table 1.

**Table 1.**
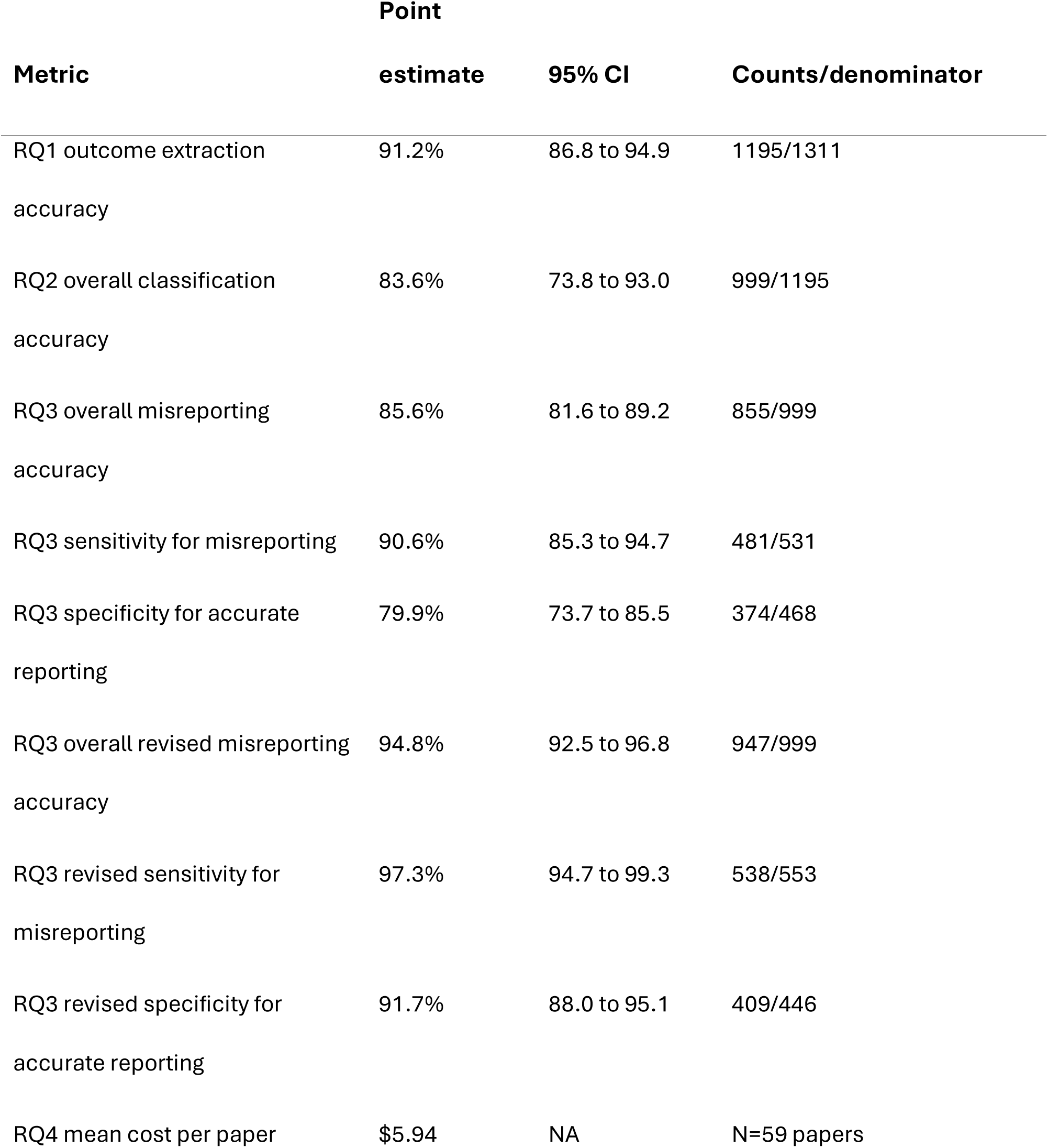
Main performance results across the central research questions for this study.

### RQ1: Outcome Extraction Accuracy

Relative to the COMPare reference standard, RegCheck achieved 91.2% recall in identifying outcomes, 95% CI [86.8, 94.9]. More specifically, in the original COMPare reports there were a total of 1,311 outcomes identified; of these, RegCheck identified 1,195 correctly, and failed to identify 116. This indicates that the automated workflow successfully recovered most of the outcomes identified through detailed manual review.

### RQ2: Outcome Classification Accuracy

Among those 1,195 outcomes present in both RegCheck and COMPare reports, RegCheck achieved 83.6% overall classification accuracy for categorising outcomes as primary, secondary, or non-prespecified, 95% CI [73.8, 93.0]. Specifically, RegCheck correctly categorized 999 of the 1,195 mutually found outcomes. The full confusion matrix for these categorisations is reported in Table 2. As well as this, per-class accuracies for primary, secondary, and non-prespecified outcomes, together with the associated counts and 95% confidence intervals, are reported in Table 3. We noted that most of the observed disagreements tended to come from protocols rather than registrations; therefore, we examined accuracies separately for the two types of document. RegCheck achieved 95.3%, 95% CI [91.4, 98.1] overall accuracy for registrations, and 69.0%, 95% CI [54.5, 86.8] overall accuracy for protocols. On further inspection, the low overall accuracy appears to originate from three specific protocols (for Trials 39, 53, and 63) with a substantial number of outcomes. RegCheck’s accuracy among the 197 total outcomes in these three protocols was 37.1%; by contrast, its accuracy among the remaining sixteen protocols’ 335 total outcomes was 87.8%.

**Table 2.** Confusion matrix for RegCheck’s outcome classification against the COMPare Trials reports for RQ2.

|  | **RegCheck:**<br>**Primary** | **RegCheck:**<br>**Secondary** | **RegCheck: Non-**<br>**prespecified** |
| --- | --- | --- | --- |
| COMPare: Primary | 83 | 2 | 4 |
| COMPare: Secondary | 9 | 589 | 161 |
| COMPare: Non-prespecified | 4 | 16 | 327 |

**Table 3.**
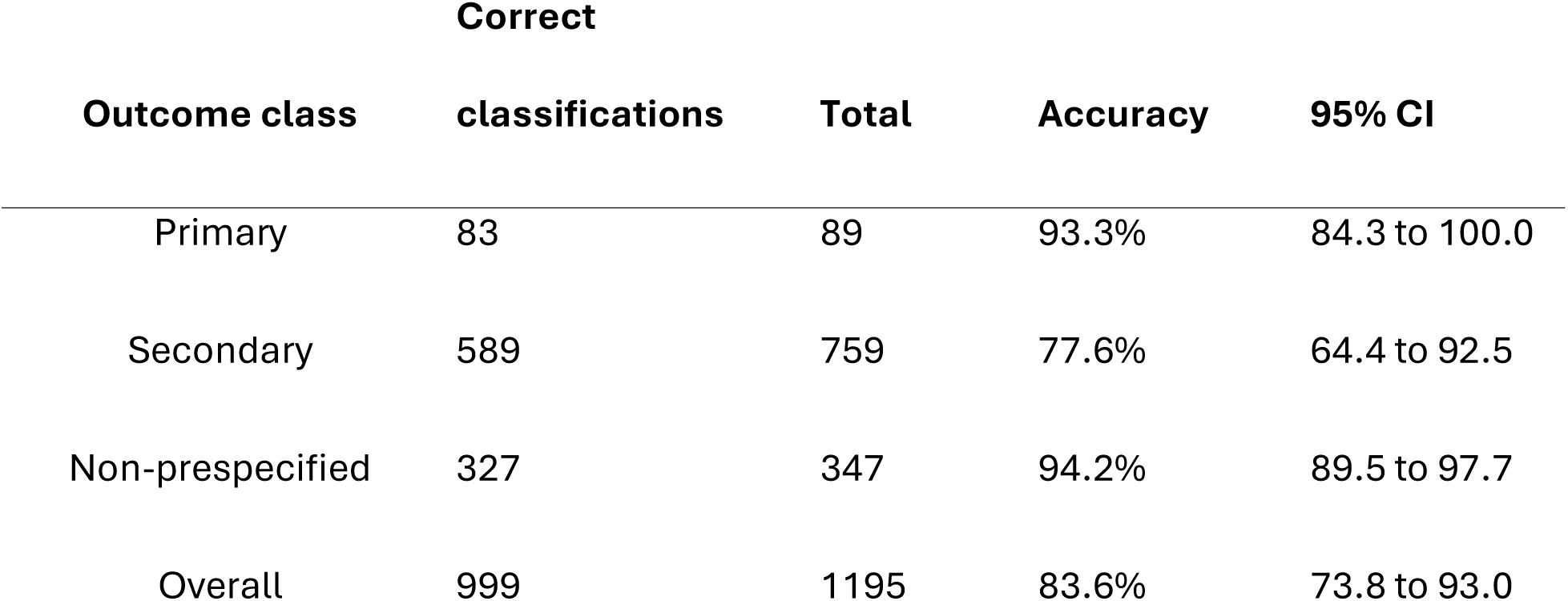
Outcome classification performance by class for RQ2.

### RQ3: Misreporting Detection Accuracy

For the detection of outcome misreporting, RegCheck achieved 85.6% overall accuracy relative to the COMPare reference standard, 95% CI [81.6, 89.2]. Sensitivity for identifying misreported outcomes was 90.6%, 95% CI [85.3, 94.7], and specificity was 79.9%, 95% CI [73.7, 85.5]. Counts underpinning these estimates are reported in Table 4 together with the full confusion matrix for these classifications.

**Table 4.**
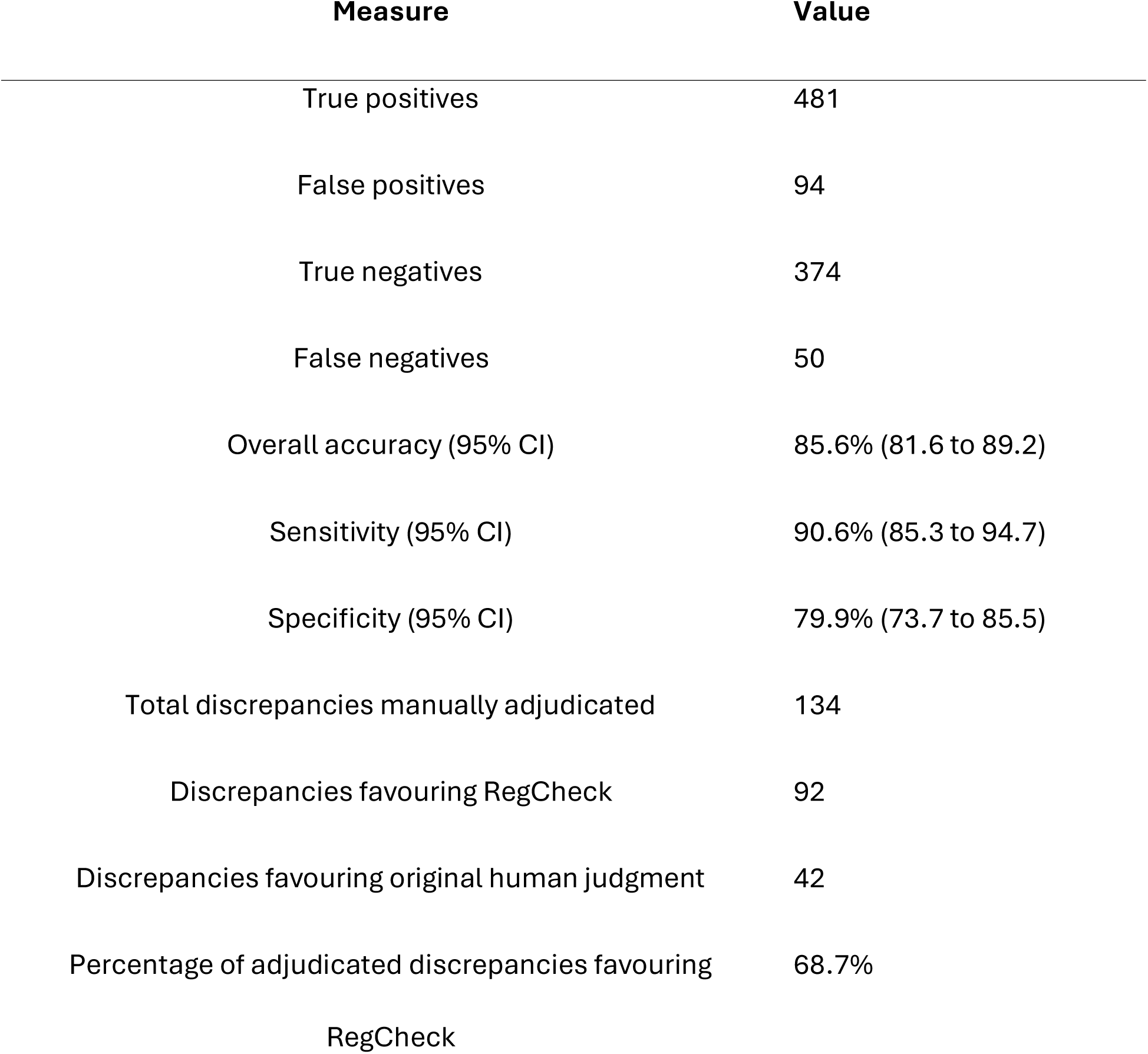
Outcome misreporting detection performance of RegCheck against the COMPare Trials reports (RQ3).

| Measure | Value |
| --- | --- |
| True positives | 481 |
| False positives | 94 |
| True negatives | 374 |
| False negatives | 50 |
| Overall accuracy (95% CI) | 85.6% (81.6 to 89.2) |
| Sensitivity (95% CI) | 90.6% (85.3 to 94.7) |
| Specificity (95% CI) | 79.9% (73.7 to 85.5) |
| Total discrepancies manually adjudicated | 134 |
| Discrepancies favouring RegCheck | 92 |
| Discrepancies favouring original human judgment | 42 |
| Percentage of adjudicated discrepancies favouring<br>RegCheck | 68.7% |

There were 144 total discrepancies between RegCheck and COMPare judgements of misreporting. We performed manual adjudication of these discrepancies in the following manner: first, the lead author (JC) reviewed all discrepancies and made manual judgements of accuracy (i.e., were RegCheck or COMPare judgements correct). Thereafter, one of two other authors (HD or NJD) reviewed these judgements and the original materials, and provided their own additional judgement. In cases where the authors disagreed, these cases were discussed until mutual agreement was found. These manual reviews of RegCheck-COMPare discrepancies suggested that 92 favoured RegCheck and 42 favoured the original human judgments. In other words, 68.7% of apparent disagreements appeared, on manual review, to reflect accurate interpretations by RegCheck rather than failures of judgement. The discrepancy adjudication summary is reported in Table 4.

Although not part of our initially registered analysis plan, it appears informative to update our original accuracy, sensitivity, and specificity estimates for RQ3 considering these manual reviews. When accounting for these resolved discrepancies, RegCheck’s updated accuracy is 94.8%, 95% CI [92.5, 96.8]; its updated sensitivity is 97.3%, 95% CI [94.7, 99.3]; its updated specificity is 91.7%, 95% CI [88.0, 95.1]. These updated values are reported in Table 1.

### RQ4: Cost

The cost of running the RegCheck workflow across all papers in the final sample (i.e., excluding the five used for piloting) was 187.59 USD for LLM-related costs, and an additional 163.96 USD for document-parsing costs (for trial protocols). As such, the total cost was 351.55 USD, which translates to an average cost of approximately 5.94 USD per paper. Although we do not have a breakdown of the specific per-paper costs, it should be noted that those papers which used protocols (rather than trial registrations) will have had a much higher average cost: partly because of the additional document-processing costs mentioned above, and partly because (for non-RAG LLM calls) there would have been a substantially larger number of tokens used within the LLM calls.

## Discussion

### Principal Findings

In this validation study, the RegCheck workflow achieved high agreement (91.2%) with the COMPare Trials benchmark for extracting outcomes, along with a lower overall rate of agreement for identifying outcome designations (83.6%); notably, accuracy for registrations (95.3%) was much higher than for protocols (69.0%). Accuracy for misreporting detection was relatively high at 85.6%. Manual adjudication of these RegCheck-COMPare disagreements suggested that most apparent disagreements reflected misjudgements in the human reference standard rather than in RegCheck’s judgement. In other words: in most cases of disagreement, RegCheck got things right that humans had got wrong. Updated accuracy for misreporting considering these resolved discrepancies (94.8%) reflected this. The workflow operated at very low marginal cost, averaging 5.94 USD per paper.

Taken together, these results suggest that automated registration-to-publication comparison of outcome reporting is technically feasible at a level likely to be practically useful. In particular, the workflow seems well-suited for use as a screening tool which may identify likely areas of concern that can then be checked more efficiently by reviewers or editors. Ideally, this tool may be useful even further back in the chain of research production: namely, for use by authors to proactively inspect their manuscript prior to journal submission to ensure that they have accurately reported all outcomes in line with CONSORT guidance.

### Interpretation

RegCheck performed well overall for outcome extraction and classification. This is encouraging because these tasks form the foundation of all downstream discrepancy assessment. If a system cannot reliably identify outcomes or their prespecified status, it cannot meaningfully assess reporting accuracy. That RegCheck performed well on both tasks suggests that this workflow can recover the structural information needed for trial auditing. The performance for outcome classification is caveated by the fact that RQ2 accuracy was substantially poorer for protocols than registrations; on inspection, this appears to largely be due to parsing failures in RegCheck’s ingestion of three specific protocols (accuracy among those three protocols: 37.1% with 197 total outcomes; accuracy among the remaining sixteen protocols: 87.8% with 335 total outcomes).

Performance was generally good for outcome misreporting identification. Notably, this task requires not only identifying corresponding outcomes across documents but also making a normative judgment about whether the reporting satisfies CONSORT-style standards of transparency. These decisions are inherently more interpretive than extraction or categorisation. Even so, the manual review of disagreements painted an interesting picture: RegCheck decisions were frequently more accurate than the original humans’ upon manual adjudication. This implies, overall, that RegCheck may function well in the role of an outcome reporting evaluator, at times rendering judgements which are more accurate than those rendered by human evaluators.

The discrepancy review also highlights an important conceptual point: when evaluating such automated approaches, it is not a reasonable expectation that these workflows may achieve 100% accuracy. After all, human evaluators making such comparisons may often disagree with one another, and “ground truth” consensus may be more difficult to reach than one may initially think. Human raters may also be conditionally influenced by previous judgements of a given paper: for instance, if a paper’s outcomes are otherwise impeccably reported, a human evaluator may be more likely to be lenient in their judgement of a single instance where an outcome is misreported. By contrast, the RegCheck workflow’s judgements of specific outcomes are made independently from other outcomes by design, preventing this unwanted variability.

### Strengths and Limitations

A major strength of this study is the use of the COMPare Trials dataset, which is one of the best known and most rigorous manual investigations of trial outcome reporting.

Additionally, the modular design of the RegCheck workflow means that separate components (extraction, classification, (mis)reporting judgement) can be run and evaluated independently from one another. This means, in principle, that researchers may for example use RegCheck’s openly-available code for outcome extraction only if they so wish, which can facilitate future research projects related to outcome extraction in the future. This follows in the lineage of other newly-developed tools aimed at facilitating the at-scale analysis and extraction of clinical trials (e.g., TrialScout, 16).

The study also has limitations. First, COMPare, although highly rigorous, was conducted on trials from 10 years ago, and may not necessarily reflect how outcomes are reported and discussed today. In addition, COMPare (and thus RegCheck) focused specifically on adherence to CONSORT 2010 guidance, which differs in some ways to current CONSORT 2025 guidance. However, the benefit of RegCheck and its open codebase is that users can trivially change the reporting standard of interest (e.g., from CONSORT 2010 to CONSORT 2025) by merely changing the content of one of its prompts. Another limitation was that the sample was restricted to a specific corpus of trials from high impact journals and may not fully represent the broader clinical trial literature. Additionally, as with all LLM-based workflows, performance estimates may vary across models, prompts, parsing quality, and document structures (17,18). As such, the results reported here are specifically applicable to the use of GPT-5.1 on high reasoning effort, and should not be generalized beyond this.

### Implications for Editors, Reviewers, and Journals

The primary practical implication of this work is that a highly-accurate system for automated examination and detection of outcome (mis)reporting is, in principle, achievable. A workflow, such as RegCheck, could in principle be integrated into pre-review editorial checks, peer review support, or post-publication monitoring (e.g., an OutcomeTracker, like the TrialsTracker project for monitoring the reporting of clinical trials data; 19,20). We would like to caveat that the deployment of such a solution should not, under any circumstances, be used in place of human judgement. Rather, RegCheck (or a similar workflow for outcome (mis)reporting) should act as a tool which can support reviewers, editors, and journals in conducting outcome reporting accuracy checks.

This is especially relevant because outcome discrepancy checking is currently rare in routine journal workflows, despite strong reporting standards and longstanding recognition of the problem. A low-cost automated screening tool could reduce the burden on editors and reviewers, increase the consistency of checks, and make it more likely that undeclared switching is identified before or after publication.

### Future Research

Future work should assess performance across wider sets of journals, registries, and trial designs; compare different model families and prompting strategies; and examine how editors and reviewers use automated discrepancy reports in practice. Future work must also tackle the ingestion issues which appear to arise in the case of a small number of protocols; or alternatively, constrain the scope of RegCheck’s use to registrations only. It will be critical to evaluate whether editors and reviewers use RegCheck in practice as a support tool (as intended), or whether they merely outsource judgements to the tool entirely. If the latter, preventative steps will need to be taken to reduce the likelihood of this occurring. It will also be important to test whether hybrid workflows, in which automation produces a draft report that is then rapidly reviewed by a human, improve both speed and accuracy relative to human-only evaluation.

### Conclusions

RegCheck exhibited high performance relative to the COMPare Trials reports in extracting outcomes, identifying their designation, and detecting outcome misreporting. After manual adjudication of RegCheck-COMPare disagreements in outcome misreporting, RegCheck’s accuracy in detecting misreporting also appeared to be as high as 94.8%. At an average cost of about 6 USD per paper, RegCheck may offer a scalable and practical tool to support more transparent and reliable clinical trial reporting.

## Declarations

### Funding

JC and ME are supported by a joint grant in the META-REP Priority Program of the German Research Foundation (#546323839) and the Swiss National Science Foundation (#100014E_225145).

### Competing interests

JC is the developer of RegCheck. All other authors declare no conflict of interest.

### Ethical approval

This study involved analysis of publicly available trial registrations, protocols, and published articles, thus no ethical approval was required.

### Data availability

The study registration, data, processing and analysis code, and RegCheck workflow code are all openly available via the Open Science Framework (https://osf.io/mxacq).

### Author contributions

Conceptualization: JC, NJD; Data curation: JC, HD, BG, NJD; Formal Analysis: JC, HD, NJD; Funding acquisition: ME; Investigation: JC; Methodology: JC, HD, ME, IH, BG, NJD; Project administration: JC; Resources: JC, HD, BG, NJD; Software: JC; Supervision: ; Validation: JC, NJD; Visualization: ; Writing – original draft: JC; Writing – review & editing: HD, ME, IH, BG, NJD

### Transparency statement

The lead author affirms that this manuscript is an honest, accurate, and transparent account of the study being reported; that no important aspects of the study have been omitted; and that any discrepancies from the study as originally planned have been explained.

^1^ An astute reader will notice an issue with this: namely, this represents an instance of “discrepant analysis”, which is known to upwardly bias estimates of a tool’s accuracy

